# Effects of food-related odors on eating behavior: A systematic review

**DOI:** 10.1101/2025.08.31.25334414

**Authors:** Jiachun Li, Xinmeng Yang, Rene de Wijk, Arianne van Eck, Sanne Boesveldt

**Affiliations:** Division of Human Nutrition and Health, Wageningen University & Research, Wageningen, The Netherlands; WUR Food and Biobased Research, Consumer Science & Intelligent Systems, Wageningen, The Netherlands; Food Quality and Design, Wageningen University & Research, Wageningen, the Netherlands

**Keywords:** Olfaction, Food cues, Eating behavior, Appetite, Food selection, Food consumption

## Abstract

Food-related odors have been examined for their influence on different stages of eating behavior. However, the existing evidence is mixed across different study designs, making it difficult to obtain an integrated understanding. This systematic review aimed to synthesize experimental evidence on how exposure to food odors impacts different stages of eating behavior, including appetite and food craving, food choice and preference, and intake, and to explore factors that may contribute to variability in findings. PubMed, Web of Science, and Scopus were searched for studies on olfactory cues, appetite, food craving, food choice, food preference, and food intake. Of 15579 records identified, 43 met the inclusion criteria. Among these, 14 focused on appetite and craving, 18 on food choice and preference, and 16 investigated food intake. The included studies were evaluated using the Joanna Briggs Institute (JBI) tool to assess their quality. The results showed distinct patterns across outcomes: sensory-specific appetite and cravings were generally enhanced by food odor exposure, whereas effects on food choice and intake were less consistent and appeared to depend on factors such as individual characteristics and the mode of odor exposure. These findings provide a clearer direction for future research on how odors influence eating behavior, with the potential to inform strategies that promote healthier diets.

## 1 Introduction

Food begins to influence human eating well before it is placed in the mouth. From the moment individuals perceive food-related sensory cues, such as seeing the golden-brown crust of freshly baked bread, hearing the sizzling sound of cooking oil, or smelling the rich aroma of sweet pastries, their physiological (Lai et al., 2021; R. D. Mattes, 1997; Morquecho-Campos, Bikker, et al., 2020; Proserpio et al., 2017) and psychological responses (Havermans, 2013; Higgs & Spetter, 2018) begin to adjust in ways that can significantly influence subsequent eating behavior. Unlike the visual system, which can be deactivated by simply closing the eyes, the olfactory system cannot be voluntarily shut off. Due to this property, individuals are often unavoidably exposed to (food-related) olfactory cues in their environment, even without conscious awareness. As a result, (food-related) odors may easily and involuntarily shape appetite, food cravings, food choice, preference and intake.

Ambient odors are perceived through orthonasal olfaction, as volatile molecules enter the body through the nose (Rozin, 1982). Because this type of perception can take place before food is placed in the mouth and consumed, orthonasal olfactory cues plays an appetitive role by triggering anticipatory responses related to eating (Boesveldt, 2017). Previous studies have shown that exposure to food odors can not only enhance general appetite, reflecting an increased overall desire to eat (Proserpio et al., 2017; Ramaekers, Boesveldt, Lakemond, et al., 2014; Zoon et al., 2016). In addition, exposure to foods odors can also elicit a craving for specific foods that are congruent with the odor. This phenomenon is known as sensory-specific appetite (SSA) (Ramaekers, Boesveldt, Gort, et al., 2014; Ramaekers, Boesveldt, Lakemond, et al., 2014; Zoon et al., 2016). For example, exposure to the smell of pizza has been shown to increase appetite for pizza more than for other foods (Ferriday & Brunstrom, 2011). However, some studies have reported no effect of food odors on general appetite (Ramaekers, Boesveldt, Gort, et al., 2014) or on SSA (Morquecho Campos, 2021; Proserpio et al., 2017), while others have even observed opposite effects, such as a decrease in appetite following odor exposure (Massolt et al., 2010).

Beyond its potential appetitive function, food odor exposure could also contribute to subsequent eating behaviors, such as food choice and food intake. Some studies have shown that food-related odors can increase the likelihood of selecting congruent food, both in responses to food images on a screen and in real food choice tasks (Chambaron et al., 2015; Gaillet-Torrent et al., 2014; Ramaekers, Boesveldt, Lakemond, et al., 2014). Other research suggests that exposure to food odors can lead to increased intake of congruent food (Koubaa & Eleuch, 2020; Proserpio et al., 2017). Despite the supportive evidence mentioned above, several others have failed to find similar effects, indicating that findings in this area remain inconsistent.

These inconsistent findings suggest that the mechanisms through which food odors influence eating behavior may be more complex than initially assumed. It is still not known how, when, and under what conditions olfactory cues affect appetite and eating behavior (e.g., for specific odor stimuli, or in particular populations). One possible explanation for the current discrepancies found in literature lies in methodological variation across studies, which includes differences in odor exposure, participant characteristics, and outcome measures. For instance, Biswas and Szocs (2019) found that the duration of odor exposure may influence behavioral outcomes: participants were likely to purchase unhealthy after a brief exposure to indulgent odor (less than 30 seconds), but they were less likely to purchase unhealthy foods when exposure to the indulgent odor was prolonged beyond two minutes. Additionally, the impact of olfactory cues may also vary across individuals, depending on factors such as gender (Koubaa & Eleuch, 2020), body mass index (BMI) (Ferriday & Brunstrom, 2008, 2011; Tetley et al., 2009), dietary restraint (Coelho et al., 2009; Fedoroff et al., 2003; Fedoroff et al., 1997; Ferriday & Brunstrom, 2008; Jansen & van den Hout, 1991), and impulsivity (Larsen et al., 2012).

Moreover, studies have used a broad range of methods and outcome measures to assess the impact of olfactory cues on eating behavior. For example, studies on food choice have used a wide range of tasks, including menu-based selection (Chambaron et al., 2015; Gaillet et al., 2013; Gaillet-Torrent et al., 2014), computerized paradigms (de Wijk et al., 2018; Ramaekers et al., 2016; Szakál et al., 2022), and real-food choice (de Wijk & Zijlstra, 2012; Mors et al., 2018; Yang et al., 2023). These variations in task design likely influence participants’ responses and may affect final outcomes.

Given the growing body of research in this field, along with inconsistent findings and methodological diversity, a comprehensive synthesis of the existing literature is therefore warranted. This systematic review aims to synthesize current evidence on the effects of food odors on eating-related outcomes, with a particular focus on three key domains: appetite, food choice and preference, and food intake. In doing so, it explicitly examines how factors such as odor characteristics, individual differences, and outcome measures may contribute to the variability in findings. By identifying these moderating factors, the review seeks to clarify the conditions under which food-related olfactory cues influence eating behavior, and to inform the design of future studies in this area.

## 2 Method

### 2.1 Search strategy and eligibility criteria

This systematic review was conducted in accordance with the PRISMA 2020 Statement (Page et al., 2021). A systematic literature search was conducted in February 2024 using PubMed, Web of Science, and Scopus to identify relevant articles published in peer-reviewed journals. The search strategy included truncated terms and Boolean operators related to food-related odors and eating behavior, applied to titles, abstracts, and keywords. The search strategy was developed using a PICOS framework. The full search strategy, including term categories and database-specific search strings, is provided in Appendix. In addition to the electronic search, snowballing was performed by manually screening the reference lists of all included studies to identify any additional potentially eligible articles.

Studies were included if they met the following criteria: (1) employed an experimental design; (2) involved healthy human participants with normal olfactory function; (3) examined the effects of food-related olfactory cues on one or more outcomes related to eating behavior, such as appetite, craving, food choice, food preference and food intake; (4) were published in peer-reviewed journals; and (5) were written in English. There were no restrictions on publication date, as long as the above criteria were met.

Studies were excluded if they: (1) involved non-human participants (e.g., animal studies) or human participants with known olfactory impairments or clinical conditions affecting eating behavior (e.g., eating disorders); (2) only used non-food-related odors or retronasal olfactory stimuli; (3) combined olfactory cues with other sensory cues; (4) did not report outcomes related to eating behavior; (5) were not peer-reviewed publications (e.g., dissertations, conference abstracts, or grey literature); or (6) were not written in English.

### 2.2 Study selection and data extraction

All records retrieved from each database were imported into EndNote, and duplicates were removed using the software. Two reviewers (JL and XY) independently screened the titles and abstracts of all retrieved articles. Irrelevant articles were excluded at this stage. Full-text screening was independently conducted by the same two reviewers. Discrepancies at any stage were resolved through discussion, with a third reviewer (SB) consulted when necessary.

Data from the included studies were extracted using structured tables. The extracted information included study characteristics (e.g., author, year), participant details (e.g., sample size, demographics), study design, stimulus characteristics (e.g. type, delivery method, exposure duration, source), outcome measures, and key findings. Data extraction was independently conducted by two reviewers (JL and XY), and any inconsistencies were resolved through discussion.

### 2.3 Quality assessment

The risk of bias and methodological quality of the included studies were assessed using the standardized critical appraisal tools developed by the Joanna Briggs Institute (JBI). Two reviewers independently conducted the assessments to ensure consistency and minimize subjective bias. Any disagreements were resolved through discussion.

### 2.4 Heterogeneity assessment and data synthesis

Due to substantial heterogeneity in study designs, olfactory stimulation methods, and outcome measures, a meta-analysis was not deemed appropriate. The results of individual studies were summarized using tabular methods. Studies were grouped by the type of eating behavior outcome (appetite and craving, food preference and choice, and food intake) and were narratively synthesized and discussed in separate sections.

## 3 Results

As shown in Fig. 1, a total of 15579 records were identified through database searching, with one additional record from other sources. After removing duplicates, 10,426 records were screened. Following full-text assessment of 110 articles, 43 studies met the inclusion criteria and were included in the review.

**Fig.1.**
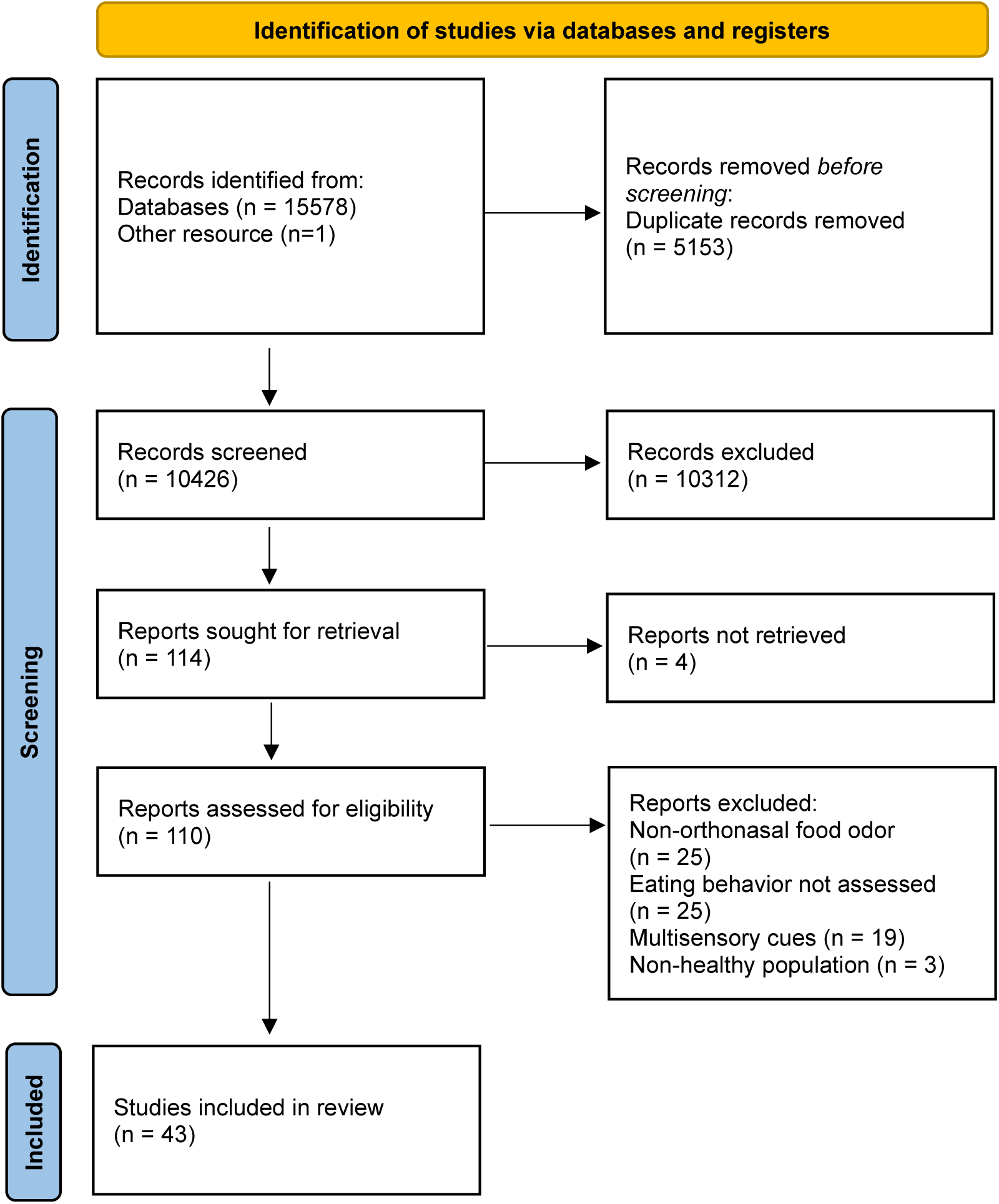
The PRISMA flow diagram.

### 3.2 Quality assessment

All 43 included studies were appraised using the JBI Critical Appraisal Checklist for Quasi-Experimental Studies. The overall methodological quality was moderate to high. Most studies clearly described their interventions, employed consistent outcome measures, and applied appropriate statistical analyses.

most studies received a “Yes” rating on seven domains. Only two domains showed greater variability. Ten studies did not have a clearly defined control group (Q2), although nine of them used valid alternative comparisons such as healthy vs. unhealthy. In addition, most studies (n = 37) either did not include pre–post measurements for all outcomes within each condition (Q5) or did so only partially. This was mainly due to the nature of certain outcomes, such as food choice or food intake, which cannot feasibly be assessed both before and after exposure. As such, the absence of repeated measures was not considered a major methodological concern in these cases.

No studies were excluded based on methodological quality. A detailed summary of the quality ratings is provided in the Appendix.

### 3.3 Outcome categories and definitions

For better synthesis, the included studies were grouped into three categories of outcomes: (i) appetite and craving, (ii) food choice and preference, and (iii) food intake.

The first category concerned appetite and craving. Appetite included both general appetite and sensory-specific appetite. General appetite refers to general desire to eat that is not specific to a particular food. Sensory-specific appetite refers to an increased desire for foods that align with the sensory cue (Ferriday & Brunstrom, 2011a; Morquecho-Campos, De Graaf, et al., 2020; Ramaekers, Boesveldt, Gort, et al., 2014; Ramaekers, Boesveldt, Lakemond, et al., 2014). None of the included studies explicitly defined craving. They simply used the term itself. Given its close conceptual relation to appetite, we classified craving within this category while retaining the terminology used by the original authors.

The second category concerned food choice and food preference. They were grouped together because both involve selecting among food options. Food choice refers to an observable decision that is linked to the possibility of obtaining the selected food. Food preference refers to comparative liking without a consequential choice, typically assessed with ranking task or repeated image-based selection tasks.

The third category concerned food intake. Food intake was defined as the amount consumed during an eating task, operationalized as grams or energy intake.

### 3.4 Effects on appetite and craving

A summary of studies investigating the impact of olfactory cues on appetite and food cravings is presented in Table 2. A total of 14 studies were included, with sample sizes ranging from 12 to 164 participants, comprising an overall total of 982 participants. Most studies (n=13) exclusively included female participants and one study included both male and female participants. Two studies explicitly categorized participants as restrained or unrestrained eaters based on restraint scores (Fedoroff et al., 2003; Fedoroff et al., 1997).

**Table 2.** Included studies on the Effects of Olfactory Cues on Appetite and Craving. The table includes details on participants, olfactory stimuli, and appetite or craving measures. Studies are ordered chronologically. In the “Relevant Findings” column, color coding indicates the direction of the effect: green = increase, red = decrease, blue = no significant change.

| Articles | Participants |  | Olfactory stimuli |  |  |  | Outcome measure |  | Relevant findings |
| --- | --- | --- | --- | --- | --- | --- | --- | --- | --- |
|  | N | Characteristics | Exposure time | Awareness of odor | Source of odor | Stimuli | Measurement timing | Appetite measure |  |
| Fedoroff et al. (1997) | 91 | - Female<br>- Restrained vs. unrestrained | 10 min | Aware | Ambient odor | Pizza<br>No odor | - Before odor exposure<br>- After odor exposure | -General appetite: hunger and desire to eat<br>- SSA: liking, desire to eat, and craving for pizza. | - General appetite was **higher** after odor exposure compared to the no-odor condition, but only among restrained eaters.<br>- SSA for pizza was **higher** after odor exposure compared to the no-odor condition, but only among restrained eaters. |
| Fedoroff et al. (2003) | 132 | - Female<br>- Restrained vs. unrestrained | 10 min | Aware | Ambient odor | Pizza<br>Chocolate-chip cookies<br>No odor | - Before odor exposure<br>- After odor exposure | SSA: liking, desire to eat, and craving for four different Foods. | - SSA for both pizza and cookies was **higher** following congruent odor exposure, compared to incongruent odor and no-odor conditions.<br>- Restrained eaters reported significantly **greater** appetite for cued food, compared to unrestrained eaters. |
| Massolt et al. (2010) | 12 | Female | 5 min | Aware | Food item | Dark chocolate<br>No odor | - Before odor exposure<br>- After odor exposure (at 5, 10, 20, 30, 40, 50 and 60 min) | General appetite: satiation, appetite, fullness, and hunger | - General appetite showed **no significant change** after chocolate odor exposure relative to pre-exposure measurements.<br>- General appetite remained unchanged in the chocolate odor group but increased in the group without odor exposure, indicating relative appetite **suppression**. |
| Styn et al. (2013) | 164 | - Female (49%) and male<br>- Smoker | 90s | Aware | Food item | Chocolate (food)<br>Staple (nonfood) | - Before odor exposure<br>- After odor exposure | Chocolate craving | Chocolate cravings **increased** significantly following chocolate odor exposure compared to pre-exposure levels. |
| Frankort et al. (2014) | 34 | Female | six times with durations of 1, 1, 2, 3, 3, and 20 minutes, respectively | Aware | Food item<br>Non-food item | Chocolate (food)<br>Pencil (non-food) | - Before odor exposure<br>- After each short odor exposure (1-3 min)<br>- During the final 20-min exposure: every 5 min | Chocolate craving | Chocolate craving **increased** after <u>short-term exposure (1-3 minutes)</u> to chocolate odor, but **decreased** following <u>prolonged exposure</u> (20 minutes), and remained unchanged in the control (non-food odor) condition. |
| Ramaekers, Boesveldt, Gort, et al. (2014) | 61 | Female | 10 min | Aware | From a cup | Natural banana<br>Artificial banana<br>No odor | - Before odor exposure<br>- During odor exposure: at 1, 5, and 9 min | - General appetite: hunger and desire to eat<br>- SSA: appetite for ‘*[specific product]*’ | - General appetite **was not affected** by smelling banana odors.<br>- SSA for banana products and sweet products **increased** following exposure to both natural and artificial banana odor, compared to no odor, whereas appetite for savory products decreased. However, SSA did not change over time. |
| Ramaekers, Boesveldt, Lakemond, et al. (2014) | 21 | Female | 20 min | Aware | Ambient odor | Chocolate (sweet)<br>Banana (sweet)<br>Meat (savory)<br>Tomato soup (savory)<br>Baked bread (staple)<br>Pine (non-food)<br>Fresh green (non-food)<br>No odor | - Before odor exposure<br>- During odor exposure: at 1, 5, and 9 min | - General appetite: hunger, desire to eat and thirst<br>- SSA: appetite for '[specific product]'? | - General appetite **increased** over time in all conditions, but food odors significantly enhanced it, while non-food odors suppressed it compared to the no-odor condition<br>- SSA for odor-specific foods **increased**, but did not change over time.<br>- Food odors had a large impact on SSA and a small impact on general appetite. |
| Firmin et al. (2016) | 92 | Female | 8s | Aware | From petri dishes | Vanilla (sweet)<br>Citrus-mint (fresh)<br>No odor | - Before odor exposure<br>- After odor exposure | Craving levels rating for each chocolate food images. | Chocolate craving **increased** following exposure to Vanilla (a sweet scent), but **decreased** after exposure to Slique Essence essential oil (a fresh scent). |
| Ramaekers et al. (2016) | 29 | Female | 5 min for each odor | Aware | From a cup | Banana (sweet)<br>Meat (savory)<br>No odor | At 1 and 5 min during the 1<sup>st</sup> odor exposure and at 7 and 11 min during the 2<sup>nd</sup> odor exposure | - General appetite: hunger and desire to eat<br>- SSA: appetite for '[specific product]' | - General appetite **was not significantly affected** by odor exposure, and **did not change** over time.<br>- SSA adjusted rapidly to the currently smelled food odor. Banana odor **increased** appetite for banana products and **decreased** appetite for meat and savory items, whereas meat odor **increased** appetite for meat products and **decreased** appetite for banana and sweet items. |
| Zoon et al. (2016) | 29 | Female | 3 min for each odor (a two-minute break between exposure) | Aware | From bottles | Chocolate (high-energy, sweet)<br>Beef (high-energy, savory)<br>Melon (low-energy, sweet)<br>Cucumber (low-energy, savory)<br>Fresh green (nonfood) | - General appetite: during the two-minute breaks between each odor exposure<br>- SSA: after all exposure | - General appetite: not mentioned details<br>- SSA: appetite for specific products | - General appetite after exposure to food odor was significantly **higher** than non-food odor and no odor.<br>- The increase in specific appetite after odor exposure was significantly **greater** for congruent foods than for incongruent or neutral foods, both in terms of taste (sweet vs. savory) and energy density. |
| Proserpio et al. (2017) | 32 | Female | 15 min for appetite (30 min in total) | Not mentioned | Ambient odor | Chocolate (high-energy, sweet)<br>Beef (high-energy, savory)<br>Melon (low-energy, sweet)<br>Cucumber (low-energy, savory)<br>No odor | - Before odor exposure<br>- During odor exposure: at 1, 8 and 15 min | SSA: desire to eat 15 different food products | - SSA for congruent food **did not change** after odor exposure.<br>- Appetite scores were **higher** during all odor conditions, regardless of the specific odor, compared to the no-odor condition |
| Wolz et al. (2017) | 39 | - Female<br>- Healthy (n=20)<br>vs. binge-eating (n=19) | 1 min | Aware | Food item<br>Non-food item | Chocolate (food)<br>Pencil (non-food) | After odor exposure | Momentary craving: desire to eat at the moment | Momentary craving **increased** significantly following chocolate odor exposure, but remained unchanged after neutral odor (pencil). |
| Morquecho-Campos et al. (2020) | 32 | Female | 3 min x 2 times | Aware | From bottles | Bread (carbohydrate)<br>Corn (carbohydrate)<br>Duck (protein)<br>Chicken (protein)<br>Butter (fat)<br>Cream (fat)<br>Cucumber (low-calorie)<br>Melon (low-calorie)<br>No odor | - Before odor exposure<br>- After the first 3-min exposure | - SSA: desire to eat '[specific product]' | - SSA for protein-rich products **increased** after exposure to protein-rich odors (duck, chicken). However, SSA was not influenced by exposure to other odor categories (carbohydrate, fat, and low-calorie). |
| Morquecho-Campos et al. (2021) | 34 | Female | 3 min x 2 times | Unaware | Ambient odor | Bread (carbohydrate)<br>Duck (protein)<br>Butter (fat)<br>Cucumber (low-calorie/control) | - General appetite: pre-exposure, 3 min post 1st exposure, 3 min post 2nd exposure.<br>- SSA: pre and post 1st 3-min exposure. | - General appetite: hunger, fullness, prospective consumption, desire to eat, and thirst<br>- SSA: appetite for '[specific product]' | - General appetite **did not change** after odor exposure compared to pre-odor exposure.<br>- SSA for macronutrient-related foods **was not significantly affected** by exposure to congruent food odors. |

Odors were administered through various methods, including ambient exposure (n=5), containers such as cups (n=2), bottles (n=2), or petri dishes (n=1), and direct presentation of food or non-food items (n=4). Exposure durations ranged from 8 seconds (Firmin et al., 2016) to 20 minutes (Ramaekers et al., 2014b). Chocolate was the most frequently used food odor (n = 8), followed by cucumber (n = 4) and banana (n = 3). In addition, five studies used non-food odors as control conditions, such as pine, fresh green, or pencil-like scents. In six studies, odors were categorized according to their taste profile (sweet vs. savory) (Proserpio et al., 2017; Ramaekers, Boesveldt, Lakemond, et al., 2014; Ramaekers et al., 2016; Zoon et al., 2016), energy density (high vs. low) (Proserpio et al., 2017; Zoon et al., 2016), or macronutrient composition (carbohydrate-, protein-, fat-based, or low-calorie) (Morquecho-Campos et al., 2020; Morquecho-Campos et al., 2021), in order to examine whether odor exposure increases appetite for foods that are congruent in sensory or nutritional properties.

The studies assessed appetite and food cravings by using different questions, which we classified into three outcome categories: general appetite, referring to appetite not specific to any particular food (n=7); sensory-specific appetite (SSA), referring to appetite for specific foods (n=8); and craving, defined as an intense desire to eat (n=4), with three studies specifically measuring chocolate craving and one assessing general craving. In some cases, both general appetite and SSA were assessed within the same study. These outcomes were based on self-reports typically collected before and after odor exposure. Five studies also included repeated assessments during exposure to capture changes over time.

Seven studies investigated changes in (self-reported) general appetite in response to food odor exposure. Two studies reported that exposure to food odors, as compared to non-food odors, increased general appetite (Ramaekers et al., 2014b; Zoon et al., 2016). Similarly, one study found that food odors increased general appetite compared to no odor, but only among restrained eaters (Fedoroff et al., 1997). Another study observed increased appetite following exposure to both food and non-food odors (Ramaekers, Boesveldt, Gort, et al., 2014). In contrast, two studies did not find a significant change in general appetite after exposure to food odors (Morquecho-Campos et al., 2021; Ramaekers et al., 2016), and one study even reported a decrease (Massolt et al., 2010).

A total of nine studies assessed sensory-specific appetite (SSA) for the odor-congruent food following exposure. About half of studies (n = 5) reported an increase in specific appetite for both the cued food and other foods within the same category (Fedoroff et al., 2003; Ramaekers, Boesveldt, Gort, et al., 2014; Ramaekers, Boesveldt, Lakemond, et al., 2014; Ramaekers et al., 2016; Zoon et al., 2016). Two studies found no significant change in SSA when comparing pre- and post-exposure to a congruent food odor (Morquecho-Campos et al., 2021; Proserpio et al., 2017). Two studies reported mixed findings. Fedoroff et al. (1997) found that the increase in SSA occurred among restrained eaters only. In another study, odors were categorized based on macronutrient content, and an increase in SSA was observed following exposure to protein-related odors (i.e. chicken and duck), but no effects were found for carbohydrate- or fat-related odors (i.e. bread and corn or butter and cream, respectively) (Morquecho-Campos et al., 2020).

Four studies explored food craving in response to odor exposure. Three studies specifically measured chocolate craving and found that brief exposure (ranging from 8 to 90 seconds) to chocolate odors (Frankort et al., 2014; Styn et al., 2013) or a sweet vanilla odor (Firmin et al., 2016) significantly increased craving for chocolate. However, Frankort et al. (2014) also reported that chocolate craving decreased following prolonged exposure (20 minutes) to the same odor. The fourth study assessed momentary craving, which refers to a person’s immediate, in-the-moment desire to eat, and found that craving ratings increased significantly in response to chocolate odor but not to a neutral odor (Wolz et al., 2017).

Overall, results suggest that food odors can variably influence appetite and craving: effects on general appetite were inconsistent across studies, sensory-specific appetite tended to increase for odor-congruent foods, and food cravings, particularly for sweet or chocolate-related items, were generally enhanced.

### 3.4 Effect on food choice and preference

A summary of studies investigating the impact of olfactory cues on food choice and preference is presented in Table 3. A total of 18 articles comprising 25 individual studies were included. Sample sizes ranged from 21 participants to approximately 900, totaling around 3000 participants. Among the 25 included studies, 19 recruited both male and female participants, while 6 studies exclusively enrolled female participants.

**Table 3.**
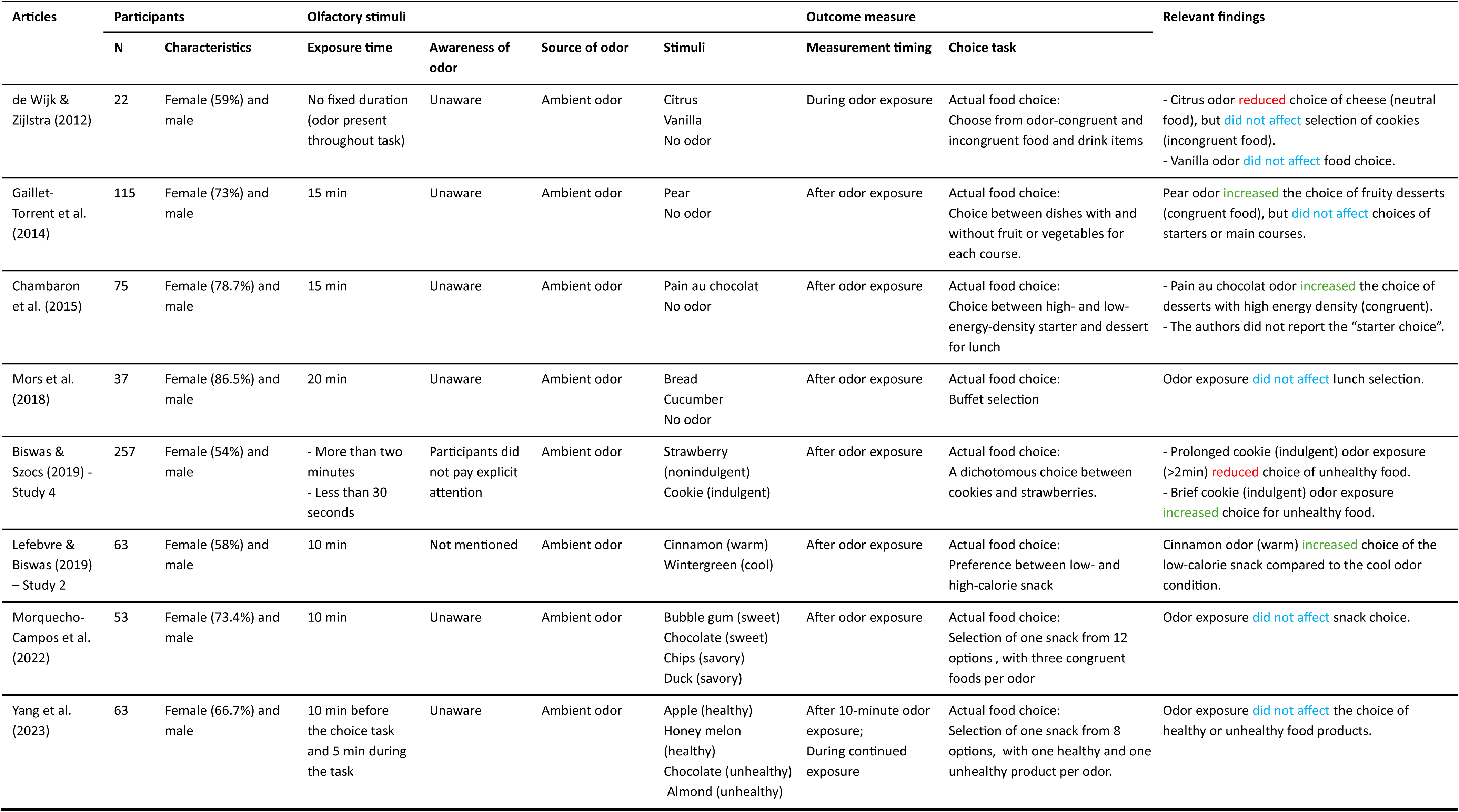
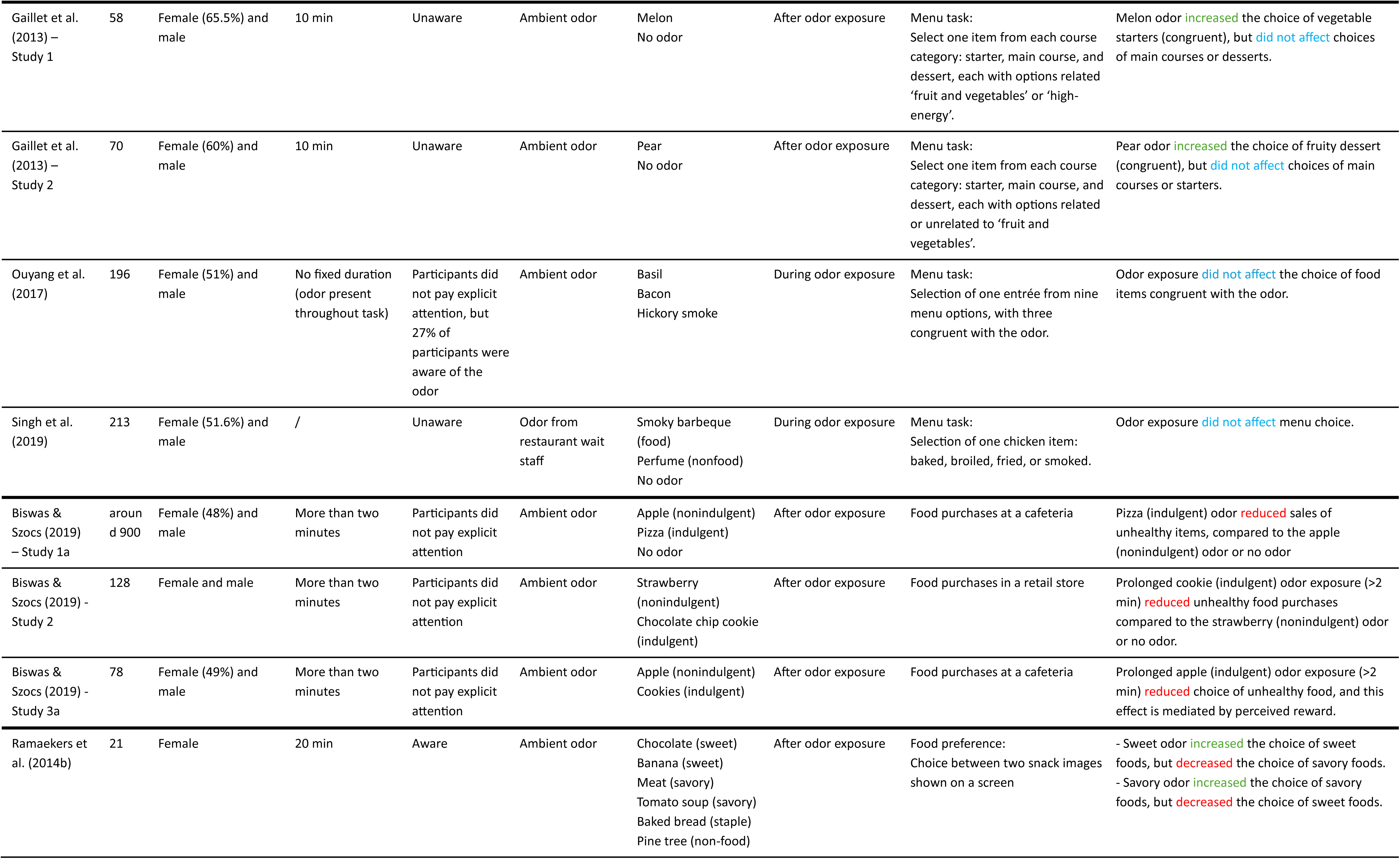
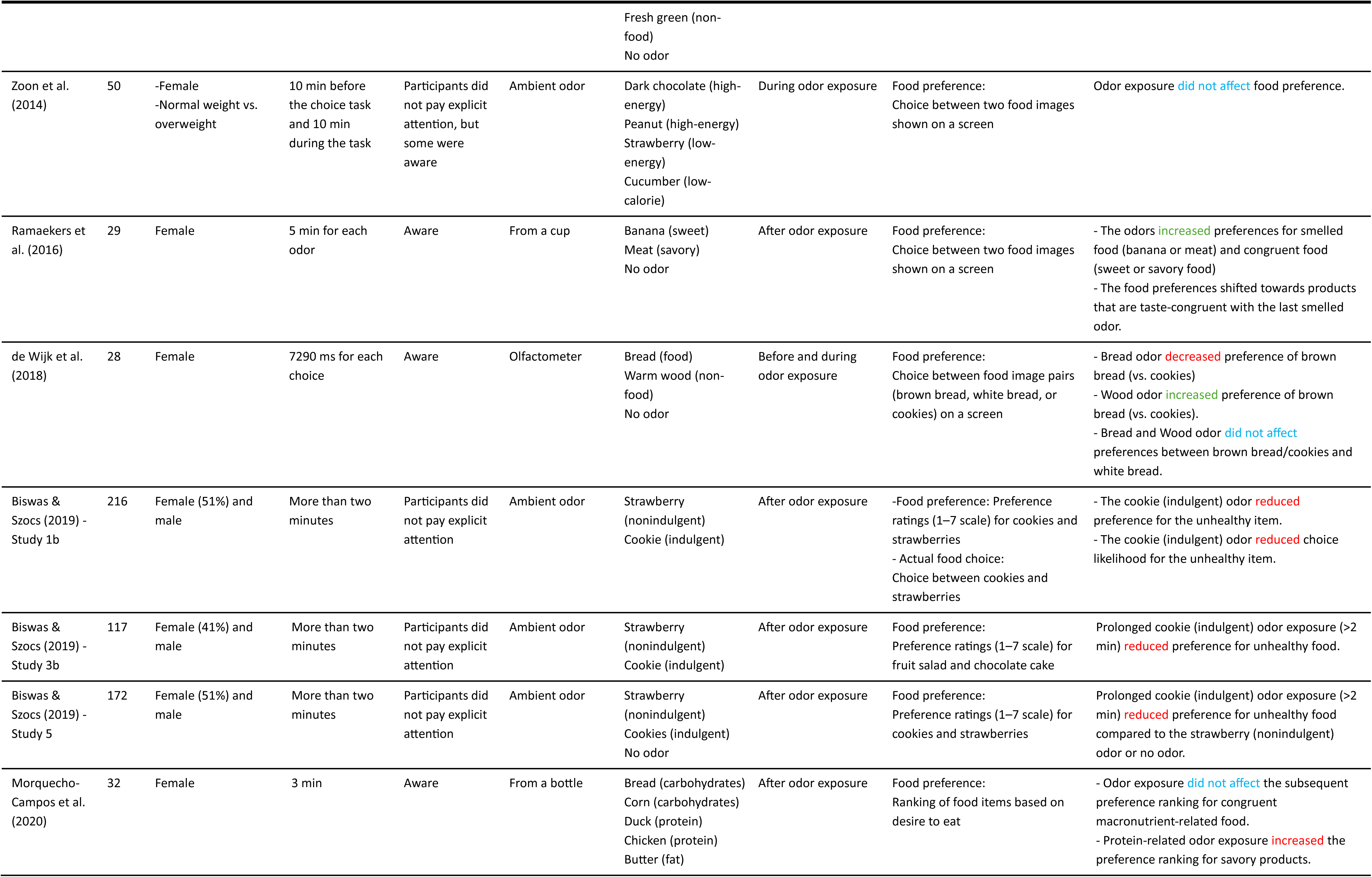
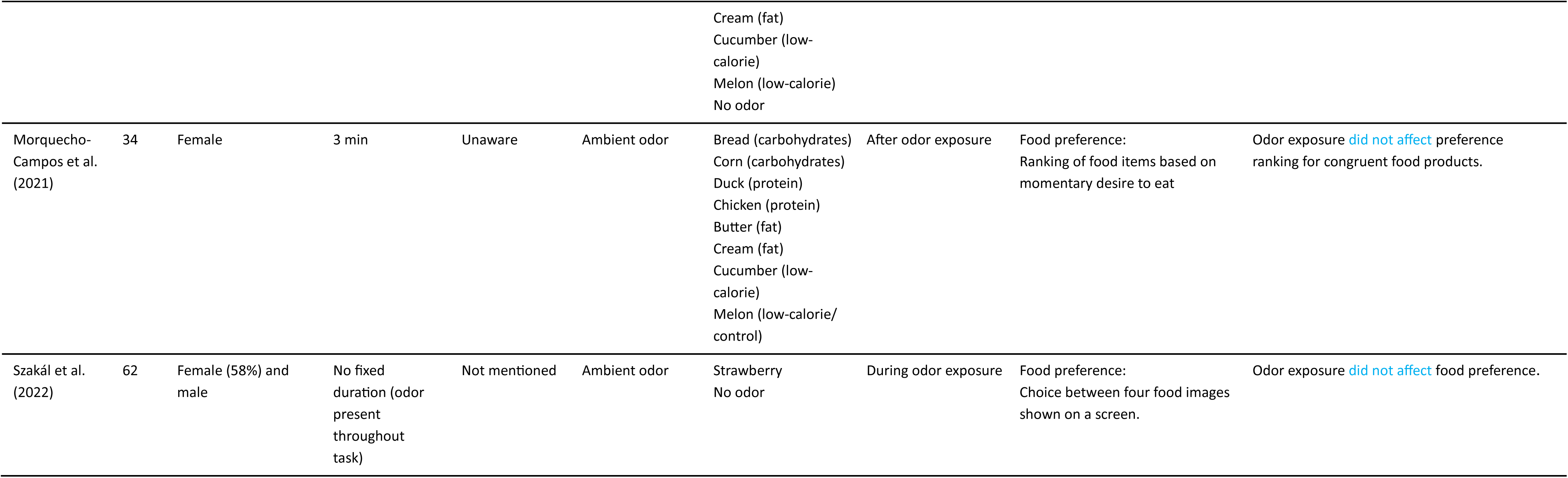
Included studies on the effects of olfactory cues on food choice and preference. The table provides details on participants, olfactory stimuli, and measures of food choice or preference. Studies are first grouped by task and then ordered chronologically within each task. In the “Relevant Findings” column, color coding indicates the direction of the effect: green = increase, red = decrease, and blue = no significant change.

Most studies (n = 22) used ambient odor. In two studies, participants were instructed to actively sniff odors from containers such as cups or bottles (Morquecho-Campos et al., 2020; Ramaekers et al., 2016), while in one study, odor was released from an odorized apron worn by a wait staff (Singh et al., 2019). Participants were typically not instructed to pay explicit attention to the odor (n=19); only the two studies involving active sniffing (Morquecho-Campos et al., 2020; Ramaekers et al., 2016) and two others required participants to consciously perceive or evaluate the olfactory cue (de Wijk et al., 2018; Ramaekers, Boesveldt, Lakemond, et al., 2014). Strawberry was the most frequently used food odor (n = 7), followed by chocolate (n = 6) and cookie (n = 6). The timing and duration of odor exposure also varied across studies. In most cases (n = 18), the odor was presented prior to the outcome measurement, after which participants completed the task in an odor-free environment. In three studies, odors were presented before the task began and remained present during the food choice task (Singh et al., 2019; Yang et al., 2023; Zoon et al., 2014). In the remaining four studies, odor exposure commenced simultaneously with the food choice task and continued until the end (de Wijk et al., 2018; de Wijk & Zijlstra, 2012; Ouyang et al., 2018; Szakál et al., 2022). Among the studies using pre-task odor exposure, durations ranged from less than 30 seconds to as long as 20 minutes. Several studies categorized odors based on specific sensory or conceptual attributes. For example, odors were classified by taste profile (sweet vs. savory), perceived healthiness (e.g., healthy vs. unhealthy or indulgent vs. non-indulgent), or thermal quality (warm vs. cool). These categorizations were used to investigate whether olfactory cues congruent with the sensory or conceptual attributes of foods would influence participants’ food choices.

The included studies employed various tasks to examine the effects of food-related odors on food choice and/or preference. Food choice tasks were used in 16 studies. Of these, nine studies asked participants to select the food they most wanted to eat among real food product, while four studies applied menu-based tasks, where participants made selections from a list of meal options (Gaillet et al., 2013: Studies 1 and 2; Ouyang et al., 2018; Singh et al., 2019). Another three field studies by Biswas & Szocs (2019) were conducted in restaurant and retail settings, assessing real-world food purchases. In addition, food preference tasks were used in ten studies. Of these, two studies used the Macronutrient Taste Preference Ranking Test/task, where participants rank-ordered multiple food items based on their desire to eat them (Morquecho-Campos et al., 2020; Morquecho-Campos et al., 2021). Three studies used a continuous forced-choice scale, asking participants to indicate their preference between two options (Biswas & Szocs, 2019: Studies 1b, 3b and 5). Five studies employed a computerized task, in which participants made repeated choices between two (de Wijk et al., 2018; Ramaekers, Boesveldt, Lakemond, et al., 2014; Ramaekers et al., 2016; Zoon et al., 2014) or four (Szakál et al., 2022) food images, indicating which item they would prefer to eat at that moment.

Of the studies examining the impact of olfactory cues on food choice, six reported no significant effect, while the remaining 10 found some influence of odor exposure. Among those showing an effect, four studies showed that food odors increased the selection of odor-congruent food items (Chambaron et al., 2015; Gaillet et al., 2013: studies 1 and 2; Gaillet-Torrent et al., 2014). However, the effects were not consistent across products. For instance, in menu-based tasks, odors increased choices of congruent starters (Gaillet et al., 2013: Study 1) or desserts (Chambaron et al., 2015; Gaillet et al., 2013: Study 2; Gaillet-Torrent et al., 2014), but had no effect on main course selections. One study reported effects on neutral food choices rather than congruent items (de Wijk & Zijlstra, 2012).

Among the ten studies assessing food preference, three showing that exposure to indulgent food scents for more than two minutes led to lower preference for the unhealthy option. Two studies used ranking tasks and categorized odors based on the macronutrient profile of the associated foods (e.g., protein-, carbohydrate-, or fat-related odors) (Morquecho-Campos et al., 2020; Morquecho-Campos et al., 2021). The study which involved conscious odor exposure via bottles found partial effects, with only protein-related odors increasing preference for savory products, while carbohydrate- and fat-related odors had no significant impact across taste or nutritional categories (Morquecho-Campos et al., 2020). In contrast, the study used unconscious ambient odor exposure and found no significant effects on food preference (Morquecho-Campos et al., 2021). Additionally, among the studies that used computerized tasks, two found that odor exposure increased preference for congruent food items (Ramaekers, Boesveldt, Lakemond, et al., 2014; Ramaekers et al., 2016), one found effects only for incongruent food preferences (de Wijk et al., 2018), and two reported no significant influence of odor cues on preference (Szakál et al., 2022; Zoon et al., 2014).

In addition, Biswas and Szocs (2019) conducted a series of experiments using both food choice and food preference tasks, and manipulated exposure duration as a moderating factor. They found that, regardless of the task, indulgent food odors increased the selection/preference of unhealthy foods when exposure was brief (under 30 seconds), but decreased when exposure was prolonged (over two minutes).

Taken together, the evidence indicates that food odors impacted food choice or preference in about half of the reviewed studies, though the direction of the effect varied across studies and was shaped by factors such as food type and exposure duration.

### 3.3 Effect on food intake

A summary of studies investigating the impact of olfactory cues on food intake is presented in Table 4. 16 studies were included, with sample sizes ranging from 32 to 213 participants and a total of 1198 participants. The majority of studies (n=13) exclusively recruited female participants, while two studies included both sexes with an approximately equal male-to-female ratio. Four studies divided participants into restrained eaters/dieters and unrestrained eaters/non-dieters based on restraint scores. One study categorized participants as high-impulsive or low-impulsive, another study grouped participants based on their tendency to eat in response to external cues (high external and low external eaters), and two study classified participants as having a normal weight or overweight based on BMI.

**Table 4.**
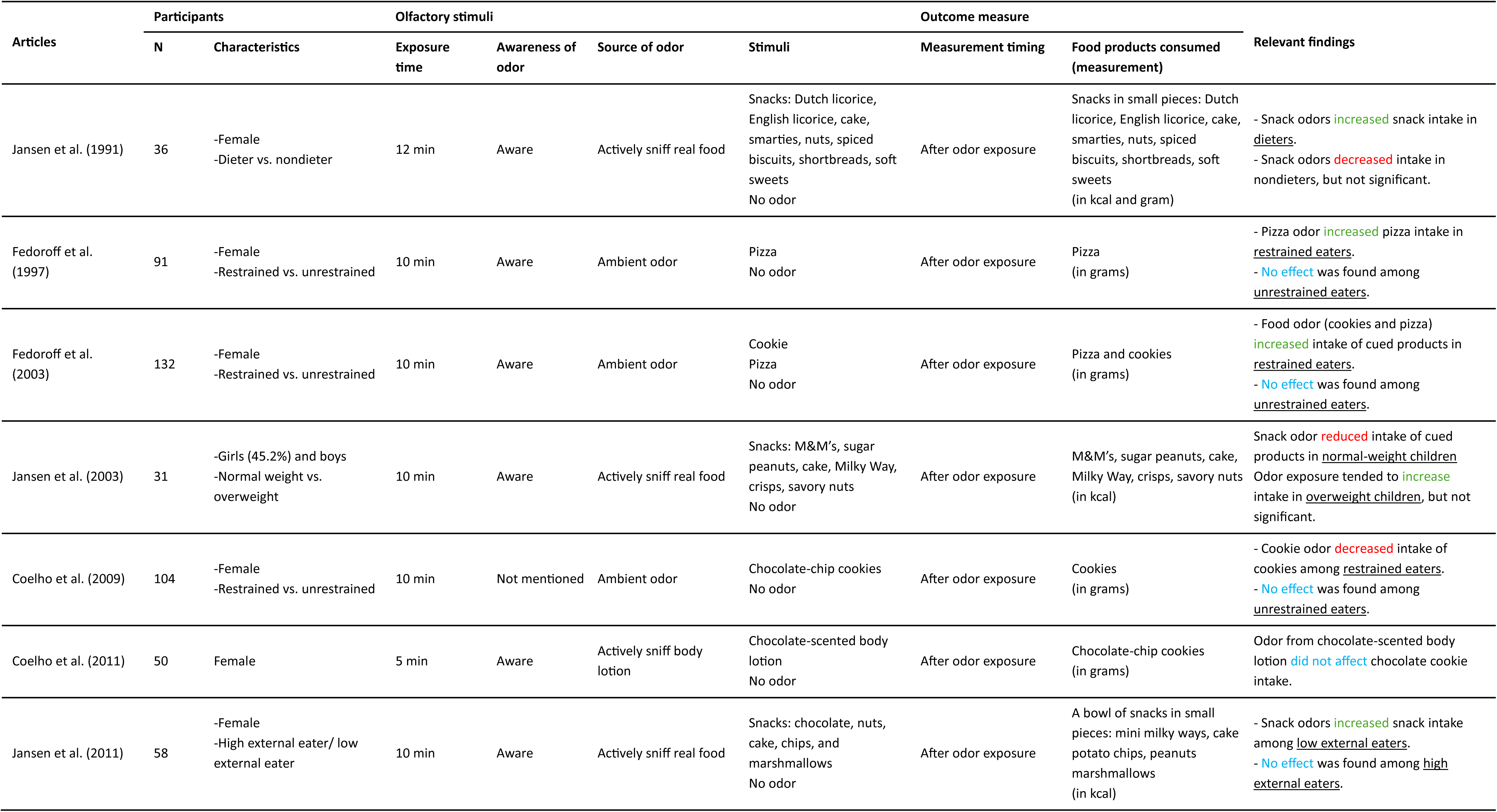
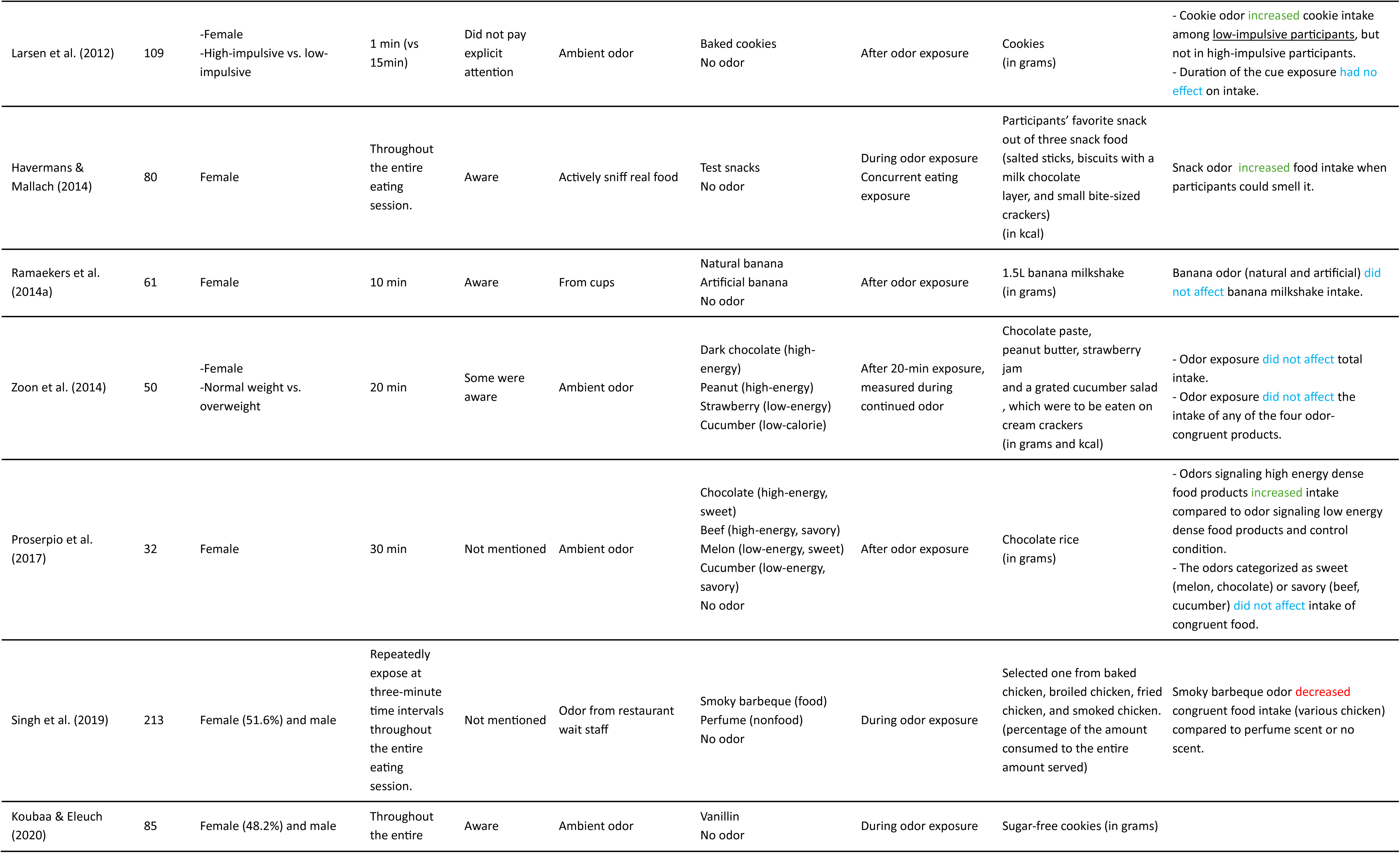
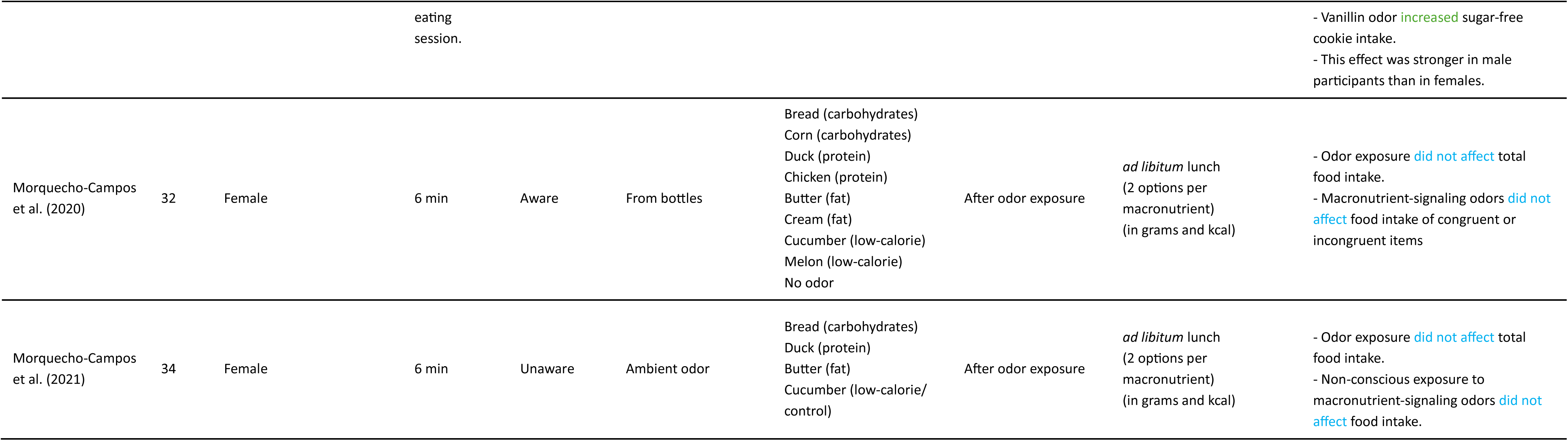
Studies on the effects of olfactory cues on food intake. The table provides details on participants, olfactory stimuli, and measures of food intake. Studies are ordered chronologically. In the “Relevant Findings” column, color coding indicates the direction of the effect: green = increase, red = decrease, and blue = no significant change. Participant characteristics are underlined.

The included studies employed a range of methods for delivering olfactory stimuli. Eight studies made use of ambient odors dispersed into the environment. In seven studies, participants actively sniffed the odors, either directly from real food or non-food items (n = 5), or from containers such as cups or bottles (n = 2). One study used an odor that was released from an odorized apron worn by a member of the restaurant wait staff. Cucumber was the most frequently used food odor (n = 4), followed by chocolate (n = 3) and cookie (n = 3). Three studies used odors derived from a mixture of snack items. In most studies (n = 13), odors were presented prior to the food intake as a form of priming, with exposure durations ranging from 1 to 30 minutes while the subsequent food intake took place in a non-odorized room. One additional study also initiated odor exposure before food intake, but the odor remained present throughout the eating episode (Zoon et al., 2014). In the remaining three studies, odor exposure occurred only during eating, and the duration varied depending on the length of the meal. One of these three studies differed in its approach: instead of adding an external odor, it manipulated olfactory input by having some participants wear a nose clip while eating, thereby eliminating odor perception to compare between the presence of natural food odor and a no-odor condition (Havermans & Mallach, 2014).

Food intake was quantified either in grams (n=9) or kilocalories (n=3), or using both kilocalories and grams (n=3) or percentage of the amount consumed (n=1). The majority of studies measured the intake of foods that were either a direct match to the olfactory cue (e.g., cookie odor followed by cookie consumption, n = 8) or belonged to the same general category as the olfactory cue (e.g., a carbohydrate-associated odor paired with the intake of carbohydrate-rich foods, n = 7). Only one study deviated from this pattern, using a sugar-free cookie that was not congruent with the presented vanillin odor (Havermans & Mallach, 2014).

Among the 16 studies included, eight reported that exposure to food odors increased the intake of congruent foods, but in five of these, the effect was observed only within specific subgroups, such as restrained eaters (Fedoroff et al., 2003; Fedoroff et al., 1997; Jansen & van den Hout, 1991), low-impulsive individuals (Larsen et al., 2012), or participants with low external eating scores (Jansen et al., 2011). In contrast, three studies reported a reduction in the intake of odor-congruent food following food odor exposure, compared to non-food or no-odor control conditions. This effect was observed in both normal-weight participants (Jansen et al., 2003; Singh et al., 2019) and among restrained eaters (Coelho et al., 2009). The remaining five studies reported no significant effect of olfactory cues on food intake.

When odors and foods were categorized by macronutrient content (Morquecho-Campos et al., 2020; Morquecho-Campos et al., 2021) or by taste quality (e.g., sweet vs. savory) (Proserpio et al., 2017), no significant effects of odor exposure on food intake were observed for congruent food items. However, when odors were categorized by energy density, one study found that exposure to high-energy odors led to significantly higher intake compared to both low-energy odors and control conditions (Proserpio et al., 2017). However, another similar study did not observe such an effect (Zoon et al., 2014). Only one study examined the effect of exposure duration, but it did not find any significant impact on food intake (Larsen et al., 2012).

Taken together, the findings suggest that odor exposure can influence eating behavior, although this effect is modulated by individual differences and tends to be weak when the food is only partially congruent with the odor cue.

## 4 Discussion

This systematic review synthesized evidence from 43 experimental studies examining how food-related olfactory cues influence various aspects of eating behavior, including appetite and craving (n = 14), food choice and preference (n = 25), and food intake (n = 16). As summarized in fig 2, findings suggest that food odors can modulate eating behavior. In particular, there was a trend toward increased SSA and craving when the odor was congruent with the food. However, effects on general appetite were more variable across studies. Evidence for effects on food choice, preference, and intake was also mixed, with outcomes seemingly moderated by individual differences (e.g., dietary restraint, BMI, impulsivity) and methodological factors such as exposure timing of odor and congruency between food and olfactory stimuli.

**Fig.2.**
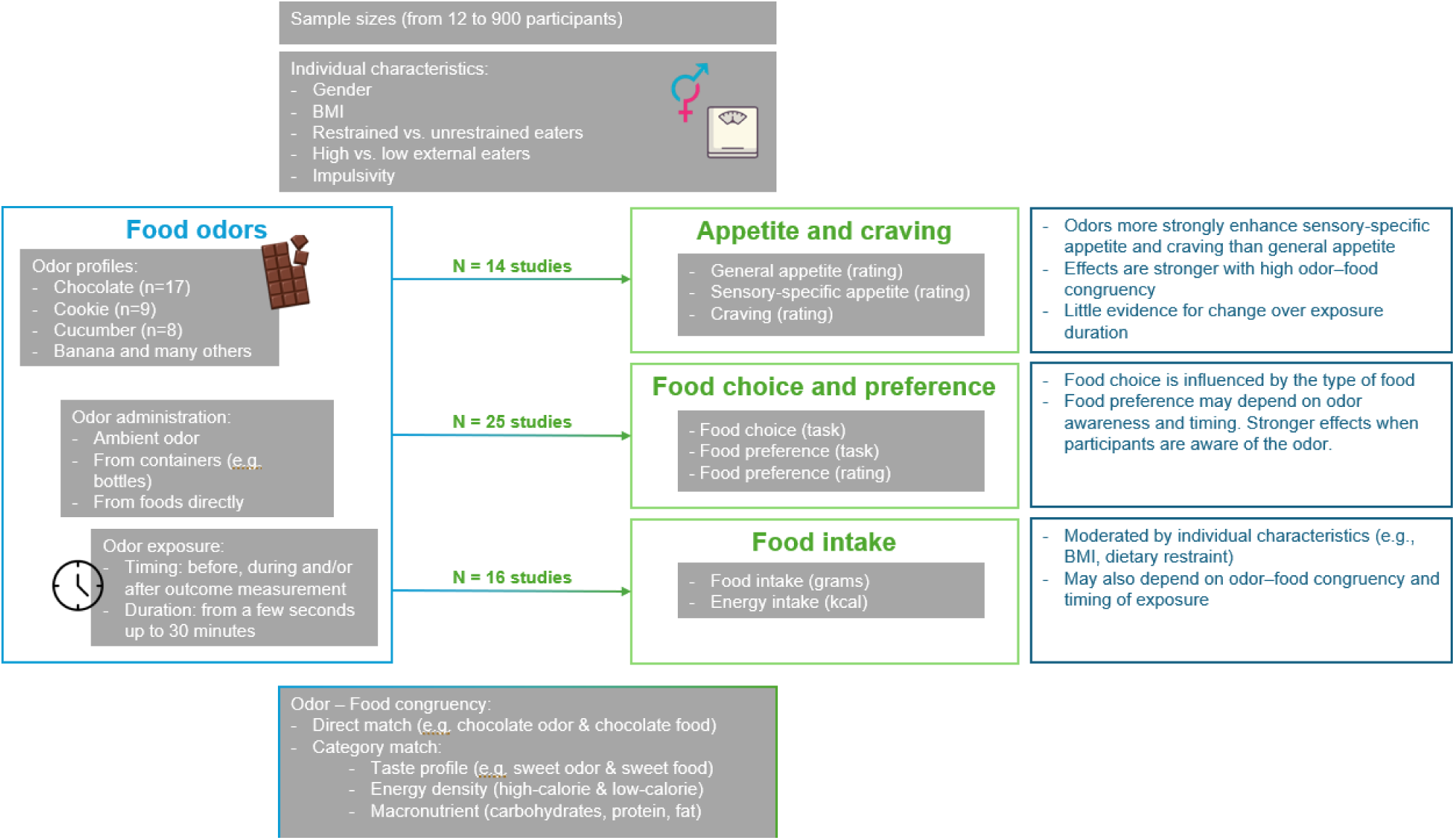
Summary of included studies, methodology, and main findings from the systematic review.

### 4.1 How food odors influence appetite and craving

Overall, the evidence suggests that while food odors do influence appetite and craving, their impact is particularly pronounced in increasing specific appetite and craving for odor-congruent foods, whereas the effect on general appetite appears to be relatively limited. This specificity effect may occur because odors allow anticipation of specific food categories, thereby selectively enhancing appetite for odor-congruent food item (Proserpio et al., 2019; Ramaekers, Boesveldt, Gort, et al., 2014). These anticipatory responses are likely shaped by learned associations between odors and the post-ingestive effects of foods, which are formed through repeated co-exposure across the lifespan (Boesveldt & De Graaf, 2017; Brunstrom & Mitchell, 2007).

Furthermore, some studies suggest that this effect is not limited to specifically matching products but can extend to broader categories of foods that share certain characteristics with the odor cue, such as taste quality (e.g., sweet vs. savory). For example, exposure to sweet odors (such as banana) has been shown to enhance appetite not only for the directly cued product (banana) but also for other sweet-tasting foods (e.g. brownie). In contrast, categorization of odors based on energy density association appears to have a less consistent influence on odor-induced appetite responses. Although some authors have proposed that odors may convey macronutrient-related information (Morquecho-Campos et al., 2019), there is currently no direct evidence that food odors influence appetite for foods within the same macronutrient category. One possible explanation is that odors used in these studies are typically those that are strongly and stably associated with a specific taste quality (e.g., chocolate and banana with sweetness, meat with salty/savoriness) (Dongen et al., 2012; Guichard et al., 2020; Teo et al., 2018), which readily elicits taste-based macronutrient associations and thus more easily increases appetite for foods within the same taste category. In comparison, the link between an odor and its associated energy density or macronutrient category is more dependent on learning and therefore less stable.

Studies measuring SSA during prolonged odor exposure generally found no significant changes over time. This suggests that even a brief odor exposure is sufficient to trigger SSA, and prolonging the exposure does not necessarily increase or reduce this effect. Only one study directly compared shorter (1–3 minutes) and longer (20 minutes) exposure durations, reporting that longer exposure actually reduced chocolate craving compared to shorter exposure (Frankort et al., 2014). For craving, however, existing studies have only used short exposure times (from 8 seconds to 3 minutes), and no evidence is currently available to determine whether this effect would change with longer exposure. Taken together, current evidence does not support a consistent effect of exposure duration on participants’ appetite and craving. Future research on food craving could introduce longer odor exposure conditions to allow comparisons of how different exposure durations affect the induction or alteration of food cravings.

### 4.2 How food odors influence food choice and preference

Although we grouped studies into two categories based on whether participants made actual food choices or only expressed subjective preferences (e.g., ratings, rankings, or image-based selections),task formats still varied considerably within each category. However, even within identical task formats, findings remained inconsistent, suggesting that task type alone does not explain the mixed results observed. We therefore considered other potential factors that might explain this variability, including the type of food, the design of the task, and exposure mode of odor.

The inconsistency in food choice outcomes may be partly explained by the type of food being selected. Odors appeared to increase the selection of congruent foods when the options involved appetizers, desserts, or snacks, whereas such effects were less frequently observed for main courses or staple foods. This is consistent with the idea proposed by Morquecho-Campos et al. (2021), who argued that the selection of main-course foods often reflects long-term habits and cultural patterns, whereas snacks and desserts are more ‘spur of the moment decisions’, easily triggered by environmental cues.

Studies utilizing food preference tasks also yielded mixed results. Preference tasks themselves varied considerably, including computerized image choices, ranking and ratings on scales. Within these computerized tasks, a notable pattern emerged from the five reviewed studies: odors presented during the task itself without participants’ explicit awareness typically failed to influence food preferences. In contrast, studies in which participants were explicitly aware of the odors and odors were presented prior to the task consistently reported significant odor-induced effects. Thus, it is likely that both odor awareness and the timing of exposure are among the factors influencing the extent to which odors affect food preferences. Conscious awareness of the odor prior to the task may lead participants to construct a distinct mental representation, which can trigger a specific appetite. This mental representation then functions as a priming effect, increasing the likelihood of choosing odor-congruent foods in later tasks. In contrast, Another potential moderator that should be considered is the duration of odor exposure. One study directly examined the effect of exposure duration on food choice and preference, finding it varied with the length of exposure. Specifically, exposure to indulgent food odors for less than 30 seconds increased the likelihood of selecting unhealthy food, whereas exposure for more than two minutes led to a decrease (Biswas & Szocs, 2019). This finding, however, should be interpreted with caution, as only limited methodological details were reported. Other studies with exposure durations exceeding two minutes did not observe similar effects, and across the studies, no clear relationship between exposure duration and outcome was found. Given the current evidence, exposure duration does not appear to serve as a moderator of the effect of odor on food choice.

Future research should therefore investigate whether the effect of exposure duration on food choice and preference emerges only under certain conditions, such as when choosing between indulgent and healthy options. It would also be valuable to examine whether the effect of exposure duration is linear or follows an inverted-U pattern, with an optimal duration beyond which the effect reverses.

### 4.3 How food odors influence food intake

The effects of odor cues on food intake are mixed across studies. These inconsistencies appear to depend on several moderating factors, such as participant characteristics and the mode of odor exposure.

Among the potential moderators, individual traits such as self-regulation, BMI, impulsivity, and external eating have been identified as possible factors influencing responses to odor cues. When participants were categorized into subgroups, the effects of odor exposure consistently differed across groups. For example, two studies found that individuals with low levels of external eating or impulsivity were more susceptible to odor-induced eating, whereas those scoring high on these traits were less affected. When participants were grouped according to BMI, the study found that that odor exposure increased intake of cued food among overweight children, while reducing among normal-weight children. Another individual factor is dietary restraint. In this review, three out of four studies found that odor exposure increased intake among restrained eaters compared with unrestrained eaters (Fedoroff et al., 2003; Fedoroff et al., 1997; Jansen & van den Hout, 1991). This may be due to the heightened sensitivity of overweight and restrained individuals to sensory cues (Ferriday & Brunstrom, 2011b; Herman & Polivy, 2008). However, one study (Coelho et al., 2009) seemingly found the opposite: restrained eaters consumed less following exposure to a food odor. The authors suggested that this reduction in intake could be explained by the Counteractive-Control Theory, which posits that when faced with temptation such as an odor cue, individuals may activate their long-term goals (e.g., weight loss) in order to suppress short-term desires (e.g., consumption of high-calorie foods). Neuroimaging data provides support for this theory, showing that exposure to tempting food cues activates not only reward-related regions but also areas involved in self-control, such as the lateral prefrontal cortex and anterior insula. This self-regulatory activation has been found to increase with personal dieting goals (Smeets et al., 2013). These contrasting findings in restrained eaters highlight the complexity of the relationship between odor exposure and food intake. It remains unclear to what extent individual differences moderate these effects. While theories such as cue sensitivity and counteractive control provide plausible explanations, further research is needed to clarify the mechanisms.

Another noteworthy pattern concerns odor–food congruency. In eight studies, participants consumed foods that exactly matched the odor cue, and they consistently reported an change on intake, either increases or decreases. However, because these effects consistently appeared in studies that also involved subgroup analyses, it remains unclear whether the observed outcomes were due to cue–food congruency or underlying participant characteristics. In contrast, in eight studies where the food was only somewhat congruent with the odor, such as when the odor and food belonged to the same energy density or taste category, significant effects on intake were observed in only three, albeit in different directions. This suggests that exact matching may be important to influence food consumption, while partial or conceptual congruency may be insufficient.

Most studies in the review used ambient, orthonasal odor exposure *before* food consumption, using it as a priming cue. Under these conditions, the majority of studies did not find an effect in healthy individuals, or reported effects only in specific subgroups. In contrast, three studies where odor exposure occurred *simultaneously* with food intake consistently reported significant effects. However, as the odors, foods, and procedures used in these studies varied considerably, the direction of the effects also differed. Two studies showed an increase in intake, whereas one reported a decrease. The study reporting decrease (Singh et al., 2019) suggests that during eating, congruent external odors may integrate with retronasal odors from the food, resulting in stronger overall olfactory stimulation that can enhance satiety and potentially reduce intake. However, in studies showing increased intake, other sensory inputs were limited, so the presence of additional odors during eating may have enhanced the overall eating experience and thereby led to greater consumption. This effect may result from odor exposure interacting with other sensory inputs, such as taste and texture, through multisensory integration.

In addition to the timing of exposure, several studies also varied the duration of odor presentation. However, across included studies, no consistent trend was found with changes in exposure duration. Only one study directly compared short (1 minute) and long (15 minutes) exposure durations and found no effect of exposure length on the intake of odor-congruent food. Therefore, the influence of pre-meal odor exposure duration on intake remains unclear. Given earlier observations suggesting that the timing of odor presentation may influence eating behavior, future research could focus on comparing odor exposure before and during eating, and then examine whether duration of exposure within these periods can enhance intake or, conversely, induce satiety and reduce food consumption.

### 4.4 Findings across outcomes

Nine studies included in this review assessed more than one outcome measure. A consistent pattern was observed between SSA and food preference: when odors increased SSA for congruent foods, participants typically also showed stronger preferences for those foods (Ramaekers et al., 2014b; Ramaekers et al., 2016). Conversely, when odors had no effect on SSA, or influenced SSA only within a specific category, preferences were either unaffected or shifted only within that same category (Morquecho-Campos et al., 2020; Morquecho-Campos et al., 2021). These findings suggest good alignment between odor-induced specific appetite and preference formation.

By contrast, food intake showed less clear association with SSA or food preference. Regardless of whether food odors induced changes in SSA or food preference, changes in intake did not consistently follow these patterns. This suggests that odor-induced appetite may play a role in shaping preference or choice, but changes in appetite or liking do not reliably predict subsequent intake. One possible explanation is that subjective reports of appetite or preference primarily reflect one’s current state or attitude toward food, which can be easily influenced by odors, whereas actual consumption is determined by a broader range of factors, including social context, physiological state, and energy needs (Boesveldt & De Graaf, 2017; Holt et al., 2017; R. Mattes, 1990).

The timing of odor exposure appears to have differential effects across stages of eating behavior. Odors presented during task performance generally did not alter specific appetite, nor did they influence preference or choice for congruent foods. In contrast, odors presented during the eating phase did affect food intake. This may be because specific appetite, food choice and preference rely more on priming effects (Boesveldt & De Graaf, 2017), whereas orthonasal odor exposure during consumption interacts with taste and retronasal aroma to alter the eating experience itself, thereby modulating the amount of food consumed (Djordjevic et al., 2004; Koubaa & Eleuch, 2020; Singh et al., 2019).

Additionally, congruency between odor cue and food appears to play a moderating role, as both appetite and food intake are more strongly affected when the odor is fully congruent with the cued food. However, when the odor and the food are only partially congruent, such as belonging to the same taste or energy category but not representing the exact same item, the effects tend to be weaker.

Finally, the effects of odors on both appetite and food intake may differ depending on dietary restraint, while other individual characteristics such as BMI, impulsivity, or external eating have shown subgroup differences primarily in food intake, with limited or no evidence of such effects on appetite or food preference.

### 4.5 Limitations in literature and future research

#### 4.5.1 Participants characteristics

Many of the included studies, particularly those investigating appetite and food intake, recruited only female participants. This gender imbalance limits the generalizability of the findings, as olfactory sensitivity and food-related responses have been shown to differ between males and females (Pfabigan et al., 2022; Sorokowski et al., 2019). In addition, some studies focused exclusively on participants from a single country or cultural background. Cultural differences may influence how individuals respond to olfactory cues and shape the types of food–odor associations they have learned, which may limit the cross-cultural applicability of the results (Ayabe-Kanamura et al., 1998). Furthermore, several studies did not report or control for individual differences in olfactory function or baseline dietary preferences. For example, a strong liking or aversion towards specific foods may confound the contribution of odor to intake, yet many studies did not control for this factor. Finally, most studies included only adult participants, with little to no data from children. Given that children’s sensory responses and eating behaviors differ from those of adults, the absence of younger age groups limits our understanding on how children respond to food-related olfactory cues.

To make research findings more reflective of real-world situations and generalizable to a broader population, future studies should, include participants of different genders and cultural backgrounds, and expand the sample to cover a wider age range, such as children and older adults. In addition, greater attention should be given to participant screening, not only assessing olfactory function but also considering habitual consumption of the target food, in order to exclude extreme cases that may bias the overall results.

#### 4.5.2 Odor presentation

The methods used to deliver odor stimuli varied considerably across studies, and some of them may lack ecological validity. For example, one study using olfactometers typically deliver odors through tubes inserted into the nostrils, which differs substantially from how odors are encountered in real-world eating situations. Results derived from such methods may not fully generalize to naturalistic food contexts. In other studies, odors were either dispersed into the ambient environment or presented in containers that participants were asked to sniff directly. The latter approach often increases participants’ awareness of the odor and engages more deliberate cognitive processing. This is especially relevant for subjective ratings like appetite, which require participants to introspectively assess their current state. In such cases, conscious awareness of the odor may increase the likelihood of associating it with internal sensations, potentially biasing the results. Moreover, the types of odors used across studies were relatively limited and often skewed toward specific categories. Commonly used odors included chocolate, cookies, cucumber, etc., with a strong emphasis on sweet-smelling stimuli. This narrow selection may restrict the generalizability of findings.

Therefore, in future research, studies should aim to increase ecological validity by using more realistic settings and odor delivery methods. For example, ambient scent diffusion in naturalistic environments, such as cafeterias, restaurants, or grocery stores, may better reflect how people encounter food-related odors in daily life. In addition, future work should also broaden the range of odors used, including savory and other everyday scents, to determine whether their effects on eating behavior are limited to the sweet odors that have been most frequently tested.

#### 4.5.3 Odor-food congruency

Previous research has classified odors and food products according to specific categories. While congruency based on certain categories, such as taste profile, appears to influence the effect, congruency based on other categories, such as macronutrient composition, shows little impact. Notably, no study to date has provided a clear and consistent definition of odor–food congruency. Therefore, further research is needed to establish a more precise conceptualization of congruency and to identify the degree or type of odor–food match that is most likely to influence the effect.

#### 4.5.4 Selective reporting bias

Selective reporting was observed in several studies. In some cases, only significant outcomes were presented, while non-significant or inconsistent results were omitted. This issue considerably limits the interpretability and integration of findings in the current review. Future research should ensure transparent reporting by specifying primary outcomes and presenting all results, regardless of statistical significance.

### 4.6 Limitations in systematic review

Finally, several limitations of this review itself should be noted. First, only studies published in English were included, which may have led to the exclusion of relevant studies reported in other languages. Second, due to the considerable heterogeneity in outcome measurements across studies, a meta-analysis was not conducted. This limits our ability to quantitatively estimate the overall strength of the association between olfactory cues and eating behaviors.

## 5 Conclusion

This review shows that food-related odors can influence multiple stages of eating behavior, but their effects differ depending on the stage and are shaped by various specific moderating factors. Odors tend to enhance sensory-specific appetite and food craving more than general appetite, particularly when the odor is highly congruent with the cued food. These effects appear to be stable over time, with little evidence for changes based on exposure duration. In the context of food choice, odor effects are influenced by factors such as the type of food involved, the individual’s awareness of the odor, and the timing of odor exposure. For food intake, the effects of odor cues appears to be moderated by participant characteristics such as dietary restraint and BMI, the degree of odor–food congruency, and whether the odor is presented before or during consumption. Overall, the impact of olfactory cues on eating behavior is context-dependent, and future research should aim to clarify the conditions under which these cues exert the strongest and most consistent effects.

## Data Availability

All data produced in the present work are contained in the manuscript

## CRediT authorship contribution statement

**Jiachun Li:** Conceptualization, Writing – original draft, Writing – review & editing, Formal analysis, Data curation, Visualization.

**Xinmeng Yang:** Writing – review & editing, Methodology, Data curation.

**Rene de Wijk:** Conceptualization, Writing – review & editing, Methodology, Supervision.

**Arianne van Eck:** Writing – review & editing, Visualization, Project administration, Methodology, Supervision.

**Sanne Boesveldt:** Conceptualization, Writing – review & editing, Project administration, Methodology, Supervision.

## Acknowledgements

This work was financially supported by the China Scholarship Council (CSC) and International Flavors & Fragrances (IFF). We would like to particularly acknowledge Parvaneh Parvin for her valuable advice on literature search and screening.

## Declaration of generative AI and AI-assisted technologies in the writing process

During the preparation of this work the authors used ChatGPT in order to the writing process to improve the readability and language of the manuscript. After using this tool/service, the authors reviewed and edited the content as needed and takes full responsibility for the content of the published article.

## Appendix Search Term Categories Used in Strategy Development

Search Term Categories Used in Strategy Development
| Concept | Subject Headings | Free-text Terms |
| --- | --- | --- |
| Population | Humans (included), Olfaction Disorders (excluded)<br>Eating Disorders | Included: Individuals,<br>Excluded: Olfaction Disorder,<br>Smell Disorders,<br>Phantosmias, Smell<br>Dysfunction, Olfactory<br>Impairment, Paraosmias,<br>Impaired Olfactions,<br>Cacosmia, Dysosmias |
| Intervention | Odorants, Cues, Smell | Cue, Odor, Odors, Food-cue,<br>Aromas, Aroma, Scent,<br>Olfaction, Olfactory,<br>Olfactory cue |
| Outcomes | Food Preferences, Choice<br>Behavior, Eating, Energy<br>Intake, Appetite, Appetite<br>Regulation, Craving, Feeding<br>Behavior | Food Preference, Food<br>Selection, Food Choice,<br>Eating Behavior, Food Intake,<br>Food Consumption,<br>Macronutrient Intake,<br>Micronutrient Intake,<br>Appetite Alterations |
| Study type | Review (excluded) |  |

### Pubmed

| Search | Query | Results |
| --- | --- | --- |
| #11 | #3 AND #6 AND #9 NOT #10 | 6,093 |
| #10 | "review"[Publication Type] | 3,268,945 |
| #9 | #7 NOT #8 | 22,997,990 |
| #8 | "Olfaction Disorders"[MeSH Terms] OR "Olfaction Disorder*"[Title/Abstract] OR "Smell Disorder*"[Title/Abstract] OR "Phantosmia*"[Title/Abstract] OR "Smell Dysfunction"[Title/Abstract] OR "Olfactory Impairment"[Title/Abstract] OR "Paraosmia*"[Title/Abstract] OR "Impaired Olfaction*"[Title/Abstract] OR "Cacosmia*"[Title/Abstract] OR "Dysosmia*"[Title/Abstract] | 7,001 |
| #7 | "Humans"[MeSH Terms] OR "Human*" [Title/Abstract] OR "Individual*" [Title/Abstract] | 23,004,289 |
| #6 | #4 OR #5 | 575,960 |
| #5 | "food preference*" [Title/Abstract] OR "eating*" [Title/Abstract] OR "food selection*" [Title/Abstract] OR "food choice*" [Title/Abstract] OR "eating behavior*" [Title/Abstract] OR "food intake*" [Title/Abstract] OR "food consumption*" [Title/Abstract] OR "feeding behavior*" [Title/Abstract] OR "macronutrient intake*" [Title/Abstract] OR "micronutrient intake*" [Title/Abstract] OR "dietary intake*" [Title/Abstract] OR "ingestion*" [Title/Abstract] OR "nutrient intake*" [Title/Abstract] OR "calorie intake*" [Title/Abstract] OR "appetite alteration*" [Title/Abstract] OR "satiety*" [Title/Abstract] OR "Craving*" [MeSH Terms] | 300,180 |
| #4 | "food preferences" [MeSH Terms] OR "choice behavior" [MeSH Terms] OR "eating" [MeSH Terms] OR "energy intake" [MeSH Terms] OR "Appetite" [MeSH Terms] OR "appetite regulation" [MeSH Terms] OR "feeding behavior" [MeSH Terms] OR "Craving" [MeSH Terms] | 365,881 |
| #3 | #1 OR #2 | 313,869 |
| #2 | "Cue" [Title/Abstract] OR "Odorant" [Title/Abstract] OR "Odor*" [Title/Abstract] OR "Odour*" [Title/Abstract] OR "Food-cue*" [Title/Abstract] OR "Aroma*" [Title/Abstract] OR "Scent*" [Title/Abstract] OR "Olfaction*" [Title/Abstract] OR "Olfactory*" [Title/Abstract] | 279,487 |
| #1 | "Odorants" [MeSH Terms] OR "Cues" [MeSH Terms] OR "Smell" [MeSH Terms] | 77,080 |

### Web of scicence

Web of science
| Search | Query | Results |
| --- | --- | --- |
| #7 | #1 AND #2 AND #5 NOT #6 | 16,271 |
| #6 | DT=("Review") | 2,679,447 |
| #5 | #9 NOT #10 | 12,720,095 |
| #4 | TS=(Olfaction Disorder* OR Smell Disorder* OR Olfactory disorder OR Olfactory dysfunction OR Phantosmias OR Smell Dysfunction OR Olfactory Impairment OR Paraosmias OR Impaired Olfactions OR Dysosmias OR Smell loss OR Cacosmia OR Dysosmia) OR TI=(Olfaction Disorder* OR Smell Disorder* OR Olfactory disorder OR Olfactory dysfunction OR Phantosmias OR Smell Dysfunction OR Olfactory Impairment OR Paraosmias OR Impaired Olfactions OR Dysosmias OR Smell loss OR Cacosmia OR Dysosmia) OR AB=( Olfaction Disorder* OR Smell Disorder* OR Olfactory disorder OR Olfactory dysfunction OR Phantosmias OR Smell Dysfunction OR Olfactory Impairment OR Paraosmias OR Impaired Olfactions OR Dysosmias OR Smell loss OR Cacosmia OR Dysosmia) | 14,582 |
| #3 | ALL=(Human* OR Human Subject* OR Participant* OR Individual* OR Male OR Female) | 12,727,594 |
| #2 | TS=(Food Preference* OR Choice Behavior OR Eating OR Energy Intake OR Appetite OR Appetite Regulation OR Feeding Behavior OR Satiety Response OR Craving OR Food Selection OR Food choice OR Food intake OR Food consumption OR Feeding Behavior OR Macronutrient Intake OR Micronutrient Intake OR Dietary Intake OR Ingestion OR Nutrient Intake OR Calorie Intake OR Energy intake OR Appetite Alterations OR Satiety OR eating behavior) | 858,737 |
| #1 | TS=(Odorant* OR Food cue OR Food cues OR Cue* OR Smell OR Odor* OR Odour* OR Aroma* OR Scent* OR Olfaction* OR Olfactory) | 745,851 |

### Scopus

| Search | Query | Results |
| --- | --- | --- |
| #7 | #1 AND #2 AND #5 AND NOT #6 | 9,743 |
| #6 | DOCTYPE ( re ) | 4,641,030 |
| #5 | #3 AND NOT #4 | 32,977,652 |
| #4 | TITLE-ABS-KEY ( “olfaction disorder*” OR “olfactory disorder” OR “smell disorder*” OR phantosmias OR “smell dysfunction” OR “olfactory dysfunction” OR “olfactory | 7,790 |
|  | impairment” OR paraosmias OR “impaired olfaction*” OR cacosmia OR dysosmias) |  |
| #3 | TITLE-ABS-KEY (human OR humans OR individual OR individuals OR Subject OR Subjects OR Participant OR Participants OR Male OR Female) | 32,984,938 |
| #2 | (TITLE-ABS-KEY ("food preference" OR "food preferences") OR TITLE-ABS-KEY (eating) OR TITLE-ABS-KEY ("eating behavior" OR "eating behaviour") OR TITLE-ABS-KEY ("energy intake") OR TITLE-ABS-KEY (appetite) OR TITLE-ABS-KEY ("feeding behavior" OR "feeding behaviour") OR TITLE-ABS-KEY (craving) OR TITLE-ABS-KEY ("satiety response") OR TITLE-ABS-KEY (satiety) OR (satiation) OR TITLE-ABS-KEY ("choice behavior" OR "choice behaviour") OR TITLE-ABS-KEY ("food selection" OR "food selections") OR TITLE-ABS-KEY ("food choice") OR TITLE-ABS-KEY ("food intake") OR TITLE-ABS-KEY ("dietary intake") OR TITLE-ABS-KEY ("food consumption") OR TITLE-ABS-KEY (ingestion) OR TITLE-ABS-KEY ("nutrient intake") OR TITLE-ABS-KEY ("calorie intake") OR TITLE-ABS-KEY ("macronutrient intake") OR TITLE-ABS-KEY ("micronutrient intake")) OR TITLE-ABS-KEY(portion size) | 891,064 |
| #1 | TITLE-ABS-KEY ( odorant ) OR TITLE-ABS-KEY ( food AND cue ) OR TITLE-ABS-KEY ( odor ) OR TITLE-ABS-KEY ( odour ) OR TITLE-ABS-KEY ( aroma ) OR TITLE-ABS-KEY ( scent ) OR TITLE-ABS-KEY ( olfaction ) OR TITLE-ABS-KEY ( "food-cue" ) OR TITLE-ABS-KEY ( olfactory ) OR TITLE-ABS-KEY(Smell) | 224,267 |

